# Phenome-wide screening of GWAS data reveals the complex causal architecture of obesity

**DOI:** 10.1101/2020.10.20.20216598

**Authors:** Luis M. García-Marín, Adrián I. Campos, Pik-Fang Kho, Nicholas G. Martin, Gabriel Cuéllar-Partida, Miguel E. Rentería

## Abstract

**Objective:** In the present study, we sought to identify causal relationships between obesity and other complex traits and conditions using a data-driven hypothesis-free approach that uses genetic data to infer causal associations.

**Methods:** We leveraged available summary-based genetic data from genome-wide association studies on 1,498 phenotypes and applied the latent causal variable method (LCV) between obesity and all traits.

**Results:** We identified 110 traits with significant causal associations with obesity. Notably, obesity influenced 26 phenotypes associated with cardiovascular diseases, 22 anthropometric measurements, nine with the musculoskeletal system, nine with behavioural or lifestyle factors including *loneliness or isolation*, six with respiratory diseases, five with body bioelectric impedances, four with psychiatric phenotypes, four related to the nervous system, four with disabilities or long-standing illness, three with the gastrointestinal system, three with use of analgesics, two with metabolic diseases, one with inflammatory response and one with the neurodevelopmental disorder *ADHD*, among others.

**Conclusions:** Our results indicate that obesity causally affects a wide range of traits and comorbid diseases, thus providing an overview of the metabolic, physiological, and neuropsychiatric impact of obesity on human health.

## INTRODUCTION

Obesity is a complex, multifactorial and preventable disease in which an imbalance between daily caloric energy intake and expenditure leads to unwanted and atypical accumulation of fat or adipose tissue, which in turn results in the impairment of human health (Hruby and Hu 2015; Purnell 2018; Panuganti et al. 2020). Obesity is the second most common cause of preventable death after smoking (Hurt et al. 2010; Mitchell et al. 2011; Ng et al. 2014; Panuganti et al. 2020), making it an essential target of public health interventions. Globally, its prevalence has increased by 27.5% for adults and 47.1% for children in the last three decades (Ng et al. 2014; Apovian 2016), affecting over 500 million adults (Panuganti et al. 2020).

Obesity is typically defined according to body mass index (BMI), which is estimated as the ratio of weight in kilograms and height in meters squared (De Lorenzo et al. 2016; Panuganti et al. 2020). Typically, an individual with obesity has a BMI higher than 30 kg/m^2^ (De Lorenzo et al. 2016; Panuganti et al. 2020). The BMI-based classification includes underweight (BMI < 18.5), normal range (18.5 < BMI < 24.9), overweight (25 < BMI < 29.9), obesity class I (30 < BMI < 34.9), obesity class II (35 < BMI < 39.9) and obesity class III (BMI > 40) (De Lorenzo et al. 2016; Panuganti et al. 2020).

Genetic epidemiological studies have made considerable advances in the understanding of the genetic propensity to obesity. Genome-wide association studies (GWAS) have identified ∼950 genomic loci associated with an obesity measure (Yengo et al. 2018; Tam et al. 2019) and genetic overlap with other anthropometric measurements, coronary artery disease, blood pressure and type 2 diabetes, among others, has been reported (Locke et al. 2015). A genetic correlation between two traits could be explained by horizontal pleiotropy (i.e. genetic variants have a direct effect on both traits) or by vertical pleiotropy (i.e. the effect of a genetic variant on a trait is mediated by its effect on another trait) (O’Connor and Price 2018; Haworth et al. 2020).

Horizontal pleiotropy represents a challenge for statistical methods seeking to determine causality between two traits. For example, traditional Mendelian randomisation (MR) methods can be confounded by horizontal pleiotropy in the presence of a genetic correlation, which increases the likelihood of false-positive findings (O’Connor and Price 2018; Koellinger and de Vlaming 2019; García-Marín et al. 2021). The Latent Causal Variable (LCV) is a recently developed statistical approach developed to investigate whether a genetic correlation between traits is explained by causal effects or by horizontal pleiotropic effects (O’Connor and Price 2018; Haworth et al. 2020; García-Marín et al. 2021).

Understanding the extent to which obesity is causally associated with other conditions is a fundamental question in obesity research. Here, we conduct a genetic screening using GWAS summary data to identify potential causal associations between obesity and other phenotypes. Specifically, we apply the LCV method to perform a hypothesis-free phenome-wide screening to the extensive collection of phenotypes with GWAS summary data (N = 1 498) compiled in the Complex Trait Genetics Virtual Lab (CTG-VL).

## METHODS

### Data

The present study used summary statistics from GWAS for obesity and 1 498 other phenotypes. Summary statistics summarise relevant parameters such as allele frequency, effect size, standard error and the p-value of genetic variants tested on the trait of interest. Several published GWAS have made available their summary statistics to the scientific community to enable researchers to advance understanding of the genetic components of several phenotypes. The CTG-VL (https://genoma.io/) (Cuéllar-Partida et al. 2019) has compiled a set of 1 610 GWAS summary statistics, and the inclusion criteria was a nominally significant heritability derived from LD-score regression. CTG-VL includes GWAS summary statistics from the UK Biobank released by Neale’s Lab (www.nealelab.is/uk-biobank/) (Neale’s Lab 2018) and from GWAS consortia. For this study, we only used GWAS derived from European populations to avoid potential biases due to population differences in linkage-disequilibrium and allele frequencies.

### Obesity dataset

The obesity GWAS summary statistics used here correspond to a sample (*N=361 194*) of European ancestry from the second wave of GWAS results released by Neale’s Lab (ICD10 code E66) (Neale’s Lab 2018; Cuéllar-Partida et al. 2019) available in the CTG-VL. Obesity was assessed as a binary trait based on BMI classification (BMI>30), and GWAS summary statistics were adjusted for age, age^2^, inferred_sex, age * inferred_sex, age^2^ * inferred_sex, and 20 genetic ancestry principal components (Neale’s Lab 2018; Cuéllar-Partida et al. 2019).

### LCV analysis

Genetic causal proportion (GCP) between the obesity GWAS and 1 498 GWAS was estimated using the phenome-wide LCV pipeline implemented in CTG-VL as described previously (Haworth et al. 2020; García-Marín et al. 2021). Briefly, GWAS summary statistics for obesity were formatted and uploaded onto CTG-VL. Then, we conducted the phenome-wide analysis pipeline (Haworth et al. 2020), which includes LD-score regression (Bulik-Sullivan et al. 2015b) as well as LCV analysis (O’Connor and Price 2018). Lastly, causal architecture plots were used to visualize the results. In particular, as part of the phenome-wide analysis pipeline, the LCV method was applied to all traits that showed a genetic correlation with obesity based on bivariate LD-score regression (Bulik-Sullivan et al. 2015b) at Benjamini-Hochberg’s False Discovery Rate (FDR < 5%). Then, to account for multiple testing on LCV estimates, we applied an FDR < 5% to the GCP estimates.

The phenome-wide analysis pipeline (Haworth et al. 2020), which is publicly available in CTG-VL, is performed in R 4.00 based on the R script that the original authors of the LCV method (O’Connor and Price 2018) have made available (https://github.com/lukejoconnor/LCV). Within this pipeline, to ensure consistency of alleles and variants across GWAS summary statistics, data is formatted using munge_sumstats.py made available by the LD-score software and extracted hapmap SNPs using the list of SNPs (w_hm3.snplist) (https://github.com/bulik/ldsc/wiki). Full details about the phenome-wide analysis pipeline in CTG-VL are described and illustrated in previous studies (Haworth et al. 2020; García-Marín et al. 2021).

The LCV method does not distinguish between the ‘exposure’ and the ‘outcome’ (O’Connor and Price 2018). These are exchangeable labels that do not affect the degree of causality; specifically, the sign of the result denotes which trait is the determinant and which trait is the outcome (O’Connor and Price 2018). Further, the LCV method estimates GCP by assuming a latent variable L that mediates the genetic correlation between two traits, which is assumed to be the causal component mediating the genetic correlation between the phenotypes (O’Connor and Price 2018; Haworth et al. 2020; García-Marín et al. 2021). The GCP ranges from −1 (full genetic causality of Trait 2 on Trait 1) to 1 (full genetic causality of Trait 1 in Trait 2). A |GCP| of 1 indicates that a genetic correlation between traits may be explained by vertical pleiotropy, whereas a GCP of 0 indicates that a genetic correlation between traits may be explained by horizontal pleiotropy. (O’Connor and Price 2018; Haworth et al. 2020; García-Marín et al. 2021). Notably, a |GCP| < 0.60 is considered low and indicates limited partial genetic causality (O’Connor and Price 2018).

### Sensitivity analysis

As a sensitivity analysis, we applied Bonferroni correction for multiple testing comparisons to identify traits with statistically significant genetic correlations and with evidence of a causal relationship with obesity based on their rG and GCP estimates. Bonferroni is a stricter, more conservative approach than FDR to determine statistical significance.

## RESULTS

We conducted a phenome-wide LCV analysis between obesity and 1 498 other phenotypes to estimate their genetic correlation and GCP. We identified 266 genetic correlations with obesity at FDR < 5%. Of those, 105 were inferred to be causally associated (|GCP| > 0.6; FDR < 5%; **Online Resource 1**) and five showed evidence of a limited partial genetic causality (|GCP| < 0.6; FDR < 5%; **Online Resource 1**). Putative outcomes of obesity included cardiovascular diseases, anthropometric measurements, the health of the musculoskeletal system, behavioural or lifestyle factors, respiratory diseases, body bioelectric impedances, psychiatric disorders, diseases of the nervous system, disabilities or long-standing illness, health of the gastrointestinal system, use of analgesics, metabolic diseases, inflammatory response and neurodevelopmental disorders, among others (**Table 1** and **Figure 1**).

**Table 1.** LCV method summary results for obesity.

| Category | Number of potential causal relationships | Number of traits influenced by obesity | Number of traits that influence obesity |
| --- | --- | --- | --- |
| Cardiovascular | 26 | 26 | 0 |
| Anthropometric measurements | 22 | 22 | 0 |
| Musculoskeletal system | 9 | 8 | 1 |
| Behavioural / Lifestyle | 9 | 9 | 0 |
| Respiratory | 6 | 6 | 0 |
| Bioelectric impedances | 5 | 5 | 0 |
| Psychiatric | 4 | 4 | 0 |
| Nervous system | 4 | 4 | 0 |
| Disabilities | 4 | 4 | 0 |
| Gastrointestinal system | 3 | 3 | 0 |
| Use of analgesics | 3 | 3 | 0 |
| Metabolic disease | 2 | 2 | 0 |
| Inflammatory response | 1 | 1 | 0 |
| Neurodevelopmental | 1 | 1 | 0 |
| Mouth problems | 1 | 1 | 0 |
| Others | 9 | 9 | 0 |

The number of potential causal relationships with obesity corresponds to those results with FDR < 5%. Obesity was tested against a panel of 1 498 potentially heritable traits in the CTG-VL catalogue.

**Fig.1.**
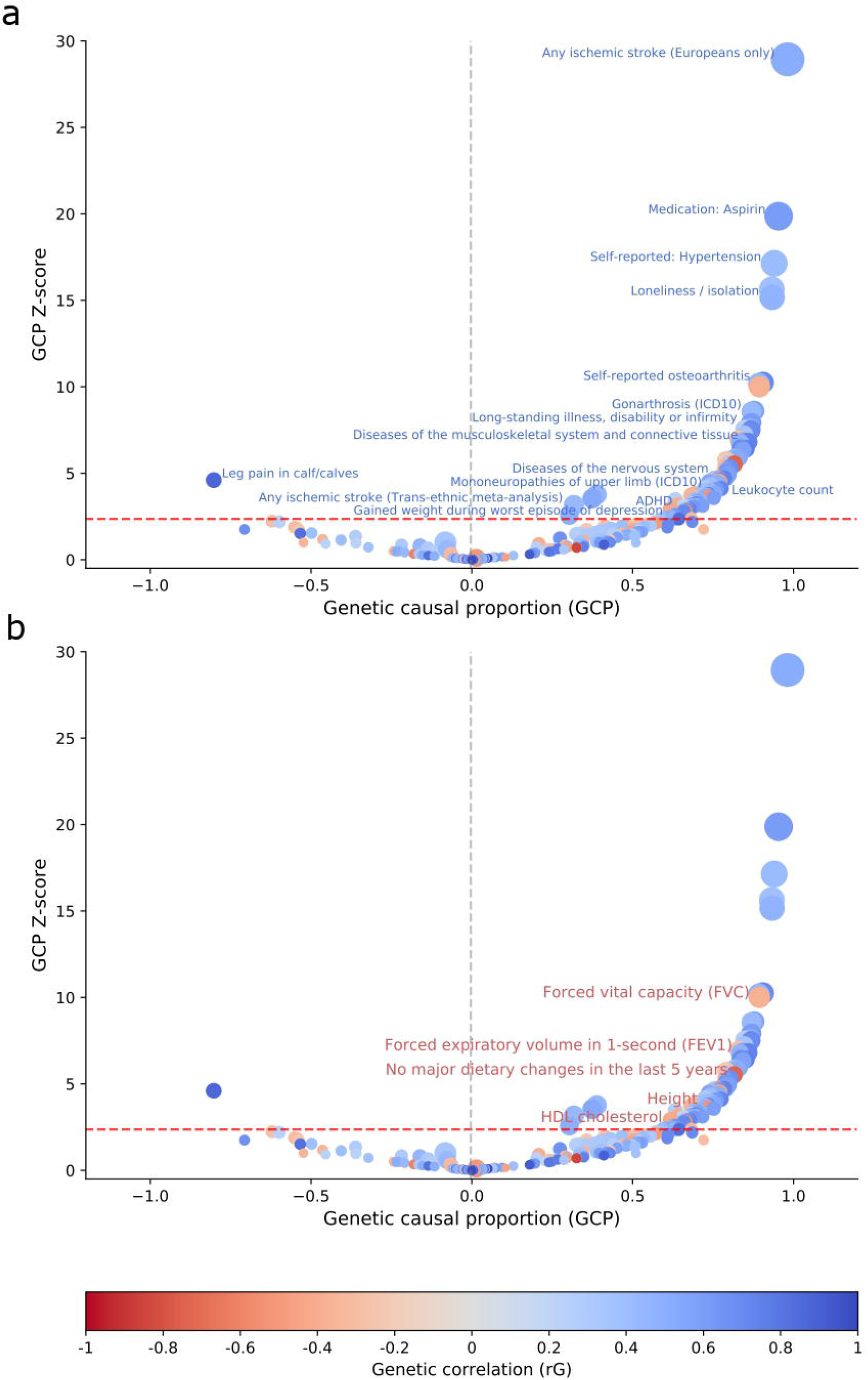
Causal associations for obesity (FDR <5%) Causal architecture plots showing the latent causal variable exposome-wide analysis results.Each dot represents a trait with a significant genetic correlation with obesity. The x-axis shows the GCP estimate, whilst the y-axis shows the genetic causality proportion (GCP) absolute Z-score (as a measure of statistical significance). The statistical significance threshold (FDR<5%) is represented by the red dashed lines, while the division for traits causally influencing obesity (on the left) and traits causally influenced by obesity (on the right) is represented by the grey dashed lines. Results are shown separately for traits with a positive genetic correlation with obesity (a) and with a negative genetic correlation with obesity (b). *Phenotypes causally associated with obesity in the present study but not with BMI in Haworth et al., 2020.

Our results show 26 cardiovascular phenotypes as potential consequences of obesity. For instance, we identified obesity as the inferred causal determinant of self-reports in *hypertension* (GCP = 0.94, p-value_GCP_ = 7.71 x 10 ^-76^) and *heart attack* (GCP = 0.39, p-value_GCP_ = 1.68 x 10 ^-04^). Conditions diagnosed by a doctor such as *high blood pressure* (GCP = 0.93, p-value_GCP_ = 4.34 x 10 ^-55^), *heart attack* (GCP = 0.32, p-value_GCP_ = 1.58 x 10 ^-03^) and *angina problems* (GCP = 0.63, p-value_GCP_ = 1.82 x 10 ^-02^) were inferred causal outcomes of obesity. A similar pattern was observed for *chronic ischaemic heart disease* (GCP = 0.72, p-value = 1.26 x 10 ^-04^) allocated in the International Classification of Diseases (ICD10) and *diastolic blood pressure*. In contrast, obesity was found to influence a decline in high-density lipoprotein (*HDL*) cholesterol (GCP = 0.68, p-value_GCP_ = 1.68 × 10^−02^; **Figure 1** and **Table 2**).

**Table 2.** Obesity is causally associated with cardiovascular, anthropometric, musculoskeletal, behavioural or lifestyle, respiratory, psychiatric, nervous system, gastrointestinal, metabolic, inflammatory response and neurodevelopmental phenotypes.

| Category | Trait | rG | GCP | GCP pval |
| --- | --- | --- | --- | --- |
| Cardiovascular | Self-reported hypertension | 0.42 | 0.94 | 7.71E-66 |
| Cardiovascular | Diastolic blood pressure | 0.27 | 0.85 | 4.97E-14 |
| Cardiovascular | *Chronic ischaemic heart disease (ICD10) | 0.41 | 0.72 | 5.94E-04 |
| Cardiovascular | Major coronary heart disease event | 0.49 | 0.74 | 1.58E-04 |
| Cardiovascular | *Myocardial infarction | 0.49 | 0.70 | 1.56E-03 |
| Cardiovascular | *HDL cholesterol | -0.32 | 0.68 | 1.68E-02 |
| Anthropometric measurement | *Height | -0.22 | 0.72 | 1.22E-04 |
| Anthropometric measurement | *Waist circumference | 0.71 | 0.66 | 5.02E-03 |
| Anthropometric measurement | Hip circumference | 0.64 | 0.63 | 9.49E-03 |
| Musculoskeletal system | Self-reported osteoarthritis | 0.71 | 0.91 | 1.20E-24 |
| Musculoskeletal system | Gonarthrosis (ICD10) | 0.57 | 0.88 | 7.16E-18 |
| Musculoskeletal system | Knee pain in the last month | 0.70 | 0.87 | 5.81E-14 |
| Musculoskeletal system | Arthrosis | 0.47 | 0.83 | 1.29E-11 |
| Musculoskeletal system | Hip pain in the last month | 0.55 | 0.79 | 2.44E-07 |
| Behavioural / Lifestyle | *Loneliness / Isolation | 0.48 | 0.93 | 6.11E-52 |
| Behavioural / Lifestyle | *Previous smoker | 0.32 | 0.72 | 3.11E-05 |
| Behavioural / Lifestyle | Previous alcohol drinker | 0.44 | 0.73 | 1.87E-04 |
| Behavioural / Lifestyle | *Age first had sexual intercourse | -0.50 | 0.77 | 2.16E-06 |
| Respiratory | *Forced vital capacity (FVC) | -0.37 | 0.89 | 1.48E-03 |
| Respiratory | Forced expiratory volume in 1 second (FEV1) | -0.27 | 0.83 | 3.69E-12 |
| Respiratory | *Shortness of breath walking on level ground | 0.76 | 0.85 | 2.16E-06 |
| Respiratory | Wheeze or whistling in the chest in the last year | 0.46 | 0.69 | 8.09E-04 |
| Psychiatric | *Ever had period of extreme irritability | 0.36 | 0.85 | 5.67E-13 |
| Psychiatric | *Manifestations of mania or irritability | 0.43 | 0.68 | 1.77E-03 |
| Psychiatric | *Gained weight during worst episode of depression | 0.70 | 0.63 | 1.82E-02 |
| Nervous system | *Diseases of the nervous system | 0.48 | 0.76 | 8.77E-06 |
| Nervous system | Mononeuropathies and upper limb (ICD10) | 0.39 | 0.72 | 1.37E-04 |
| Nervous system | Carpal tunnel syndrome | 0.43 | 0.68 | 1.27E-03 |
| Nervous system | Nerve, nerve root and plexus disorders | 0.51 | 0.66 | 3.70E-03 |
| Gastrointestinal system | *Diverticular disease of intestine (ICD10) | 0.51 | 0.67 | 3.43E-03 |
| Gastrointestinal system | *Self-reported gastro-oesophageal reflux / gastric reflux | 0.50 | 0.62 | 1.22E-02 |
| Metabolic disease | *Diabetes diagnosed by a doctor | 0.60 | 0.75 | 3.49E-04 |
| Inflammatory response | *Leukocyte count | 0.20 | 0.66 | 2.00E-03 |
| Neurodevelopmental | *Attention-Deficit - Hyperactivity Disorder (ADHD) | 0.85 | 0.64 | 1.84E-02 |

This table shows some traits with a significant (FDR<5%) strong genetic causal proportion (GCP > 0.60) for obesity. The number of potential causal relationships corresponds to those results with FDR < 5%. Due to space restrictions, all nominally significant genetic correlations for obesity are reported in **Online Resource 1**.
Category = Category of trait, Trait = Trait causally associated with obesity, rG = Genetic correlation, GCP = Genetic causal proportion, GCP pval = Genetic causal proportion unadjusted p-value.
\*Phenotypes causally associated with obesity in the present study but not with BMI in Haworth et al., 2020.

Our results show a potential causal effect of obesity on 22 anthropometric measurements. In particular, obesity influenced increments of 18 traits, including *fat percentages* throughout the body and *ankle spacing width*, while a decline in *height* (GCP = 0.72, p-value_GCP_ = 1.22 x 10 ^-04^) and *handgrip strength* were found to be causally influenced by obesity. Similarly, we found evidence of obesity influencing the decrease of five body bioelectrical impedance measures, including in both arms and legs.

Obesity was the inferred causal determinant of eight phenotypes involving *diseases of the musculoskeletal and connective tissue* (**Table 2**). Pain-related phenotypes such as *knee* (GCP = 0.87, p-value_GCP_ = 5.81 x 10 ^-14^) and *hip* (GCP = 0.79, p-value _GCP_= 2.44 x 10 ^-07^) *pain in the last month along with self-reported osteoarthritis, arthrosis and gonarthrosis (ICD10)*, were found to be causally influenced by obesity. In contrast, *leg pain in calves* (GCP = −0.80, p-value_GCP_ = 4.18 x 10 ^-06^) was an inferred causal determinant of obesity.

Six inferred causal relationships were observed between obesity and respiratory-related phenotypes. For instance, obesity was observed to increase *shortness of breath* (GCP = 0.85, p-value_GCP_ = 1.43 x 10 ^-10^) and *whistling in the chest* (GCP = 0.69, p-value_GCP_ = 8.09 x 10 ^-04^). Consistently, obesity was found to causally influence the decline of *forced vital capacity (FVC*; GCP = 0.89, p-value_GCP_ = 1.48 x 10 ^-23^) and *forced expiratory volume in one second (FEV1*; GCP = 0.83, p-value_GCP_ = 3.69 x 10 ^-12^) (**Figure 1** and **Table 2**).

Obesity was an inferred causal determinant of four psychiatric-related traits (**Figure 1** and **Table 2**), including *gaining weight during the worst period of depression* (GCP = 0.63, p-value_GCP_ = 1.82 x 10 ^-02^). Also, obesity was observed to increase irritability through traits such as *ever having a period of extreme irritability* (GCP = 0.85, p-value_GCP_ = 5.67 x 10 ^-13^), experiencing *manifestations of mania or irritability* (GCP = 0.68, p-value_GCP_ = 1.77 x 10 ^-03^) and *ever highly irritable for two days*. Similarly, an increase in the behavioural trait *loneliness or isolation* (GCP = 0.93, p-value_GCP_ = 6.11 x 10 ^-52^) was found to be caused by obesity (**Figure 1** and **Table 2**).

Obesity was an inferred causal determinant of four *diseases of the nervous system* (GCP = 0.76, p-value_GCP_ = 8.77 x 10 ^-06^; **Table 2**) including *mononeuropathies of upper limb (ICD10;* GCP = 0.72, p-value_GCP_ = 1.37 x 10 ^-04^), *carpal tunnel syndrome* (GCP = 0.68, p-value_GCP_ = 1.27 x 10 ^-03^) and *nerve, nerve root and plexus disorders* (GCP = 0.66, p-value_GCP_ = 3.7 x 10 ^-03^; **Figure 1**).

Three phenotypes related to the gastrointestinal system, including *diverticular disease of the intestine (ICD10;* GCP = 0.67, p-value_GCP_ = 3.43 x 10 ^-03^) and *self-reported gastro-oesophageal reflux* (GCP = 0.62, p-value_GCP_ = 1.22 x 10 ^-02^) were identified as potential outcomes of obesity (**Table 2**). *Diabetes*, both *self-reported* (GCP = 0.72, p-value_GCP_ = 1.90 x 10 ^-03^) and *diagnosed by a doctor* (GCP = 0.75, p-value_GCP_ = 3.49 x 10 ^-04^), showed evidence of being inferred causal outcomes of obesity. Also, obesity was also found to pose a causal effect on *high leukocyte levels* (GCP = 0.66, p-value_GCP_ = 2.00 x 10 ^-03^) and *attention-deficit / hyperactivity disorder (ADHD;* GCP = 0.64, p-value_GCP_ = 1.84 x 10 ^-02^).

### Sensitivity analysis

As a sensitivity analysis, we applied a Bonferroni correction to our results instead of FDR to account for multiple testing (Bonferroni < 0.05; Figure 2). Bonferroni is well-known to be a much more conservative approach than FDR (Noble 2009). Using this approach, 86 genetic correlations, of which 52 were inferred to be outcomes causally associated with obesity (|GCP| > 0.6; **Online Resource 2**,), were statistically significant, including anthropometric measurements, bioelectrical impedance, poor health of the musculoskeletal system, hypertension, diabetes, and ADHD (**Table 3**).

**Fig.2.** Causal associations for obesity (Bonferroni <5%) Causal architecture plots showing the latent causal variable exposome-wide sensitivity analysis results.Each dot represents a trait with a significant genetic correlation with obesity. The x-axis shows the GCP estimate, whilst the y-axis shows the genetic causality proportion (GCP) absolute Z-score (as a measure of statistical significance). The statistical significance threshold (Bonferroni<5%) is represented by the red dashed lines, while the division for traits causally influencing obesity (on the left) and traits causally influenced by obesity (on the right) is represented by the grey dashed lines. Results are shown separately for traits with a positive genetic correlation with obesity (a) and with a negative genetic correlation with obesity (b)

**Table 3.** Causal associations with obesity in the sensitivity analysis (Bonferroni < 5%).

| Category | Trait | rG | GCP | GCP pval |
| --- | --- | --- | --- | --- |
| Behavioural / Lifestyle | Loneliness / isolation | 0.48 | 0.93 | 6.11E-52 |
| Behavioural / Lifestyle | Rent accommodation | 0.61 | 0.87 | 2.32E-15 |
| Behavioural / Lifestyle | No major dietary changes in the last 5 years | -0.73 | 0.82 | 3.17E-08 |
| Behavioural / Lifestyle | Age first had sexual intercourse | -0.50 | 0.77 | 2.16E-06 |
| Cardiovascular | Self-reported: Hypertension | 0.42 | 0.94 | 7.71E-66 |
| Cardiovascular | High blood pressure diagnosed by doctor | 0.39 | 0.93 | 4.34E-55 |
| Cardiovascular | Diseases of the circulatory system | 0.59 | 0.80 | 8.77E-07 |
| Cardiovascular | HDL cholesterol | -0.32 | 0.68 | 1.68E-02 |
| Disabilities | Long-standing illness, disability or infirmity | 0.61 | 0.86 | 4.59E-15 |
| Disabilities | Blue badge disability allowance | 0.69 | 0.86 | 2.01E-13 |
| Disabilities | Disability living allowance | 0.66 | 0.80 | 3.41E-07 |
| Metabolic disease | Diabetes diagnosed by doctor | 0.60 | 0.75 | 3.49E-04 |
| Metabolic disease | Self-reported diabetes | 0.60 | 0.72 | 1.90E-03 |
| Musculoskeletal system | Self-reported osteoarthritis | 0.71 | 0.91 | 1.20E-24 |
| Musculoskeletal system | Gonarthrosis (ICD10) | 0.57 | 0.88 | 7.16E-18 |
| Musculoskeletal system | Knee pain in last month | 0.70 | 0.87 | 5.81E-14 |
| Musculoskeletal system | Diseases of the musculoskeletal system and connective tissue | 0.57 | 0.84 | 7.95E-12 |
| Musculoskeletal system | Other joint disorders | 0.63 | 0.76 | 4.82E-05 |
| Neurodevelopmental | ADHD | 0.85 | 0.64 | 1.84E-02 |
| Respiratory | Forced vital capacity (FVC) | -0.37 | 0.89 | 1.48E-23 |
| Respiratory | Shortness of breath walking on level ground | 0.76 | 0.85 | 1.43E-10 |
| Respiratory | Wheeze or whistling in the chest in last year | 0.46 | 0.69 | 8.09E-04 |
| Behavioural / Lifestyle | Loneliness / isolation | 0.48 | 0.93 | 6.11E-52 |
| Behavioural / Lifestyle | Rent accommodation | 0.61 | 0.87 | 2.32E-15 |

## DISCUSSION

In this work, we performed a phenome-wide screening of potential causes and effects of obesity on a number of health conditions. Although previous research has made extensive efforts to describe how obesity and metabolic syndrome influence several body systems, their relationships with inflammatory response, hypertension, cardiovascular disease, neurodevelopmental disorders, and the musculoskeletal and nervous system has not been fully elucidated. In this study, we found that obesity was causally associated with increased *leukocyte count, self-reported hypertension, high blood pressure and diabetes diagnosed by a doctor*. Leukocytes are white blood cells involved in both local and general inflammatory response (Langer and Chavakis 2009; Leick et al. 2014). Further, it has previously been reported that obesity increases adipose tissue dysfunction, leading to a pro-inflammatory state, which in turn can result in vascular dysfunction impairing endothelium vasodilation with an impact on hypertension and affecting the responsiveness of the insulin-vasodilator mechanism (Campia et al. 2012; Swarup et al. 2020). Also, obesity is considered the main cause of metabolic syndrome components such as high blood pressure and triglycerides, while the increased risk of diabetes is attributed to a decrease in insulin secretion as a consequence of obesity-related effects (Goodarzi 2018).

Obesity and metabolic syndrome are major risk factors for cardiovascular disease (Goodarzi 2018; Swarup et al. 2020; Panuganti et al. 2020). In the present study, several cardiovascular diseases were found to be causally influenced by obesity. Specifically, our results indicate that obesity is an inferred causal determinant of *major coronary heart disease events, heart attacks, myocardial infarctions, chronic ischaemic heart disease (ICD10) and angina problems*. Consistently, an increase in *Aspirin’s* intake, which is commonly prescribed for secondary prevention of cardiovascular diseases (Ansa et al. 2019), was also identified as an outcome of obesity. Further, it has been previously shown that inflammation is a significant risk factor for heart disease events (Hoffman et al. 2004; Kim et al. 2017). Thus, our findings support the hypothesis in which the relationship between obesity and cardiovascular diseases is mediated by inflammation from high leukocyte levels due to obesity, leading to high *blood pressure and hypertension*, which are known risk factors for cardiovascular disease. Similarly, the increase in the use of *Aspirin* could be mediated by the development of cardiovascular disease.

Obesity influences the increase in the mechanical load across weight-bearing joints, which has been associated with musculoskeletal deterioration and neuropathic pain (Anandacoomarasamy et al. 2008; Hozumi et al. 2016). Further, previous studies have suggested that an increase in fat mass may result in a decrease in bone mass (Anandacoomarasamy et al. 2008).Our findings uncovered inferred causal associations in which obesity is the putative causal determinant of musculoskeletal pain and diseases such as *osteoarthritis*. A similar pattern was observed for *diseases of the nervous system, including mononeuropathies and nerve, nerve root and plexus disorders*. Consistently, obesity was found to influence an increase in the *use of analgesics* such as *Aspirin, Codamol and Paracetamol*. Our results suggest that an increase in analgesics use may be mediated by the development of musculoskeletal pain and damage to the nervous system. It is possible that the inflammatory state induced by obesity may also result in poorer musculoskeletal and nervous system health. However, more research is needed to disentangle the complex relationships between specific-tissue inflammation, pain and obesity.

Previous studies have described an increased incidence of disability among people with obesity (Anandacoomarasamy et al. 2008; Queirós et al. 2015). Our results identified inferred causal associations between obesity and disability-related phenotypes such as *disability living allowance and long-standing illness or disability*. Thus, it is possible that the development of disabling conditions is mediated by increases in body fat, which result in poor musculoskeletal and nervous system health, which leads to a decrease in quality of life.

Despite extensive efforts to advance our understanding of the relationship between obesity and lung function, the effects of obesity on the respiratory system have not been fully elucidated. Previous findings suggest that obesity may be associated with complex respiratory diseases such as chronic obstructive pulmonary disease (COPD) and its severity (Zammit et al. 2010; Mafort et al. 2016; Dixon and Peters 2018). Consistently, our results revealed a causal effect of obesity on an increment of *shortness of breath* and *whistling in the chest*, as well as decreases in *FVC* and *FEV1*. Moreover, although our results did not identify a direct inferred causal relationship between obesity and COPD, traits such as *COPD onset and other obstructive pulmonary disease (ICD10)* were identified as causally associated with obesity. Previous studies have also reported leukocyte accumulation in lung tissue in individuals with a chronic obstructive pulmonary disease, which in turn increases the expression of adhesion molecules in bronchial blood vessels (Davis et al. 2012; Koo et al. 2017). Our results would suggest that obesity poses a deteriorative effect on the respiratory system, which could contribute to the development of complex respiratory diseases. We speculate that this relationship could be mediated by the accumulation of adipose tissue and inflammation arising from an increase in *trunk fat mass*, which in turn could lead to physiological changes decreasing lung capacity and weakening the respiratory muscles. However, more research is needed to disentangle the intricate relationship between obesity, inflammation, lung function and disease.

Observational studies have reported an association between obesity and gastrointestinal diseases such as diverticular diseases and gastro-oesophageal reflux (Camilleri et al. 2017). Similarly, increased adipose tissue has been inversely linked with adiponectin levels, which are a protective factor of gastro-oesophageal reflux complications (Chang and Friedenberg 2014). Consistently, our findings show inferred causal associations in which obesity is an inferred causal determinant of *diverticular diseases of the intestine (ICD10) and self-reported gastro-oesophageal reflux*.

Previous studies have sought to describe the extent to which obesity could be involved in the development of depression, leading to an unclear set of conclusions (Mannan et al. 2016; Day et al. 2018; Chauvet-Gelinier et al. 2019; Speed et al. 2019). For instance, some studies suggest a bidirectional relationship between obesity and depression (Mannan et al. 2016; Chauvet-Gelinier et al. 2019), while others report that anthropometric measurements such as BMI, fat mass and height are only risk factors for depression (Speed et al. 2019). Also, a Mendelian randomisation analysis reports a one-way causal association for BMI causing loneliness and a bi-directional causal association between BMI and depressive symptoms, suggesting that the relationship between these traits is complex and perhaps a consequence of shared biological mechanisms (Day et al. 2018) or lifestyle factors such as sleep quality, diet and physical inactivity (Hawkley and Cacioppo 2010). In the present study, we provide evidence for obesity as an inferred causal determinant of psychiatric-related phenotypes, such as *gaining weight during the worst period of depression* and *loneliness or isolation*. Therefore, our results suggest that obesity may contribute to an increased risk for depression, which in turn is most likely to partially mediate the association with loneliness. However, it may also be possible that there are bidirectional effects between obesity and depression (Luppino et al. 2010). As we discuss in the limitations below, LCV is not able to estimate bidirectional causality, and more research is required to investigate the extent to which depression might mediate the relationship between obesity and loneliness.

Previous research has pointed out an association between ADHD and obesity. However, cause-effect links and the underpinning molecular mechanisms of this association remain unclear (Cortese and Tessari 2017; Cortese 2019). Observational studies have examined the relationship between ADHD and obesity with both traits as exposure and outcome, suggesting that this association is independent of potential confounding factors (Cortese and Tessari 2017). Also, some genetic studies show a one-way causal relationship in which high BMI is a causal determinant of ADHD (Martins-Silva et al. 2019), while others suggest that a plausible bidirectional causal association may exist between obesity and ADHD (Liu et al. 2020). In the present study, results show an inferred causal association in which obesity is an inferred causal determinant of ADHD. However, to fully understand the effect of obesity on ADHD, additional research should seek to elucidate potential mechanisms underlying this association.

Pleiotropic effects among obesity-related phenotypes have become a focal point of interest in genetic epidemiological studies, and pleiotropy has been identified between abdominal obesity and immune pathways (Kaur et al. 2019). For instance, it has been reported that abdominal obesity increases the risk to develop autoimmune diseases due to a chronic inflammatory state (Kaur et al. 2019). Our results add up to the evidence (Lumeng and Saltiel 2011; Andersen et al. 2016; Kaur et al. 2019) suggesting that obesity prompts an immune response and a chronic inflammatory state with detrimental effects on the overall health. Certainly, the present study shows the complexity of investigating vertical (not horizontal) pleiotropic relationships, where a third phenotype could act as a mediator between the other two. For example, the association between loneliness and obesity could be partially mediated by depression. While we cannot rule out a potential causal effect of obesity on loneliness, the most likely explanation is that a shared genetic component between obesity, loneliness, and depression drives this association. Future improvements in statistical genetics methods could disentangle potential confounding effects by using causal architecture networks.

The main strengths of the LCV method as compared to traditional MR methods include (O’Connor and Price 2018; Koellinger and de Vlaming 2019; Haworth et al. 2020; García-Marín et al. 2021): (i) it is less prone to bias due to horizontal pleiotropy; (ii) it is robust to sample overlap, (iii) it uses aggregated information across the entire genome, increasing statistical power and enabling analyses between pairs of phenotypes that would be considered “underpowered” for other statistical methods.

Our results are consistent with previous studies reporting that LCV is a meaningful tool to detect potential causal associations in underpowered phenotypes for which MR methods have not been able to determine potential causation (O’Connor and Price 2018; Haworth et al. 2020; García-Marín et al. 2021). We suggest that the inferred causal relationships pointed out through LCV could be used as a testable hypothesis for future epidemiological observational and genetic studies.

Our study highlights the importance of triangulating between multiple study designs, which in turn contributes to elucidate our understanding of the complexity of obesity. Therefore, methods and findings in the present study must be compared to those in Haworth et al. 2020. The main difference between the work by Haworth et al. 2020 and ours is the design of the GWAS involved. For instance, the GWAS summary statistics used by Haworth et al. 2020 represent BMI as a continuous variable, whereas, in the present study, GWAS summary statistics correspond to dichotomised BMI based on its well-established classification for obesity (De Lorenzo et al. 2016; Panuganti et al. 2020). This difference in the study design is directly reflected in the genetic correlation between the GWAS used here and the one used by Haworth et al., 2020 (rG = 0.67, s.e. = 0.11, p-value = 7.87×10^−10^), showing that a continuous BMI measurement does not entirely reflect obesity as a disease. Still, with a well-established classification, BMI can be a useful proxy phenotype for obesity. In fact, BMI is the most commonly used proxy for obesity in large-scale public health studies (Ghesmaty Sangachin et al. 2018). Furthermore, the study by Haworth et al. 2020, assessed BMI against 1,389 other phenotypes. In contrast, in the present study, we have expanded the panel of traits and tested obesity against 1,498 other phenotypes.

Regarding the results, one of the principal differences is that in our study, we only identified one potential causal association in which obesity was the outcome of another phenotype (*leg pain in calves*), which is most likely explained by lack of physical activity. In contrast, the study by Haworth et al., 2020 identified 23 traits potentially causing BMI variation. Of those, most of them included occupational-related phenotypes, which in turn are most likely explained by lack of physical activity and socioeconomic variables such as assortative mating and educational attainment. In addition, Haworth et al. 2020 identified 110 traits as outcomes of BMI variation, whereas our study identified 109 traits as outcomes of obesity; however, a total of 68 potential causal associations were different between studies. Some of the potential causal associations that were only observed in the present study include those between obesity and *leukocyte levels, ADHD, loneliness, long-standing illness or disability, self-reported* and *diagnosed by a doctor diabetes*, cardiovascular diseases, *diseases of the nervous system*, and *diseases of the musculoskeletal system and connective tissue*, among others (**Figure 1, Table 2** and **Online Resource 1**). We attribute these substantial differences among the findings of both studies to the design of the GWAS involved.

Establishing causal associations should always arise only after convergent evidence from studies with multiple designs. Ideally, at least one should be an interventional design (e.g. a randomised controlled trial). However, interventional studies are not only expensive and time-consuming, but in many instances, are unfeasible or unethical to conduct (i.e. when an exposure known to harm participants is evaluated). In these cases, approximations using genetics to assess causality might be the best option available. Nonetheless, some limitations of the present study must be acknowledged. Although our data included GWAS summary statistics derived from consortia, which include only individuals of European ancestry but are on participants from a number of countries, most of our data was retrieved primarily from the UK Biobank, which predominantly consists of participants of European ancestry, and previous studies have highlighted ethnic differences in obesity (Higgins et al. 2019). Thus, our results’ generalizability is limited to European ancestry individuals until tested in other ethnicities.

Differences between methods used to correct for multiple comparisons should be noted. FDR has been used in previous studies describing LCV analyses results (O’Connor and Price 2018; Haworth et al. 2020; García-Marín et al. 2021). A main advantage of FDR is that it does not require tests to be independent of each other, and thus, it is useful when assessing several hypotheses that are simultaneously tested (Chen et al. 2017), like in the present study. However, FDR is less stringent than other multiple testing correction methods, such as Bonferroni (Chen et al. 2017). Although a Bonferroni correction for multiple tests is much stricter and less prone to false-positive findings, it assumes that all tests must be independent of each other (Stevens et al. 2017). This condition is not met in the present study because some GWAS included in LCV analyses are correlated (i.e., cardiovascular phenotypes, anthropometric traits, psychiatric phenotypes, among others). Here, we included results for potential causal associations between obesity and 1 498 other phenotypes using FDR < 5% correction and, as a sensitivity analysis, we have included the results for the phenome-wide analysis pipeline using a Bonferroni < 5% correction; however, we note that our tests are not entirely independent from one another.

Regarding potential bias in our analyses due to sample overlap, the LCV method and genetic correlations estimated with LD-score regression can handle sample overlap (Bulik-Sullivan et al. 2015a; O’Connor and Price 2018). Furthermore, our analyses included more than 1,400 phenotypes; however, causal associations with other traits not tested here may exist. Related to this is the interpretability of some of the phenotypes used here, such as *taking medication: Aspirin* which could be considered a proxy trait for pain or cardiovascular disease, *taking medication: Candesartan cilexetil and lisinopril* which could be considered a proxy trait for hypertension. Unfortunately, other traits for which GWAS summary statistics are available lack such a straightforward proxy interpretation.

Similarly, diabetes phenotypes, both self-reported and diagnosed by a doctor, included gestational diabetes, type 1 diabetes, type 2 diabetes, diabetes insipidus and unclassified diabetes. Considering that the physiopathology of these phenotypes is substantially different (Skyler et al. 2017), future studies should aim to further assess the associations between obesity and specific types of diabetes. In addition, even though the LCV method uses genetic information aggregated across the entire genome, the GCP estimates are still tied to the statistical power of the GWAS. Thus, the ability to infer causal associations for some phenotypes is limited, particularly for those with small sample sizes. Also, the LCV may estimate spurious associations when the genetic correlation between traits is mediated by multiple latent factors (O’Connor and Price 2018). However, the presence of multiple latent factors would reduce statistical power and lower GCP estimates biasing results towards the null (O’Connor and Price 2018). Lastly, the LCV method seeks to detect the predominant causal direction between two phenotypes (O’Connor and Price 2018; Haworth et al. 2020), and therefore, bidirectional causality cannot be tested between traits. This limitation is intrinsic to the nature of the LCV method in which a bidirectional causal association would mimic horizontal pleiotropy biasing the GCP towards the null. In our study, null findings for which a bidirectional causal hypothesis exists should be taken with caution and should be further explored in future studies using methods that can test for bidirectionality.

In summary, we assessed potential causal relationships between obesity and 1498 phenotypes and identified 110 traits with significant causal associations with obesity. Our findings uncovered the effect of obesity on leukocyte-related inflammation, which may incur in a chronic proinflammatory state and several metabolic syndrome components. Further, we provide evidence for the impact of obesity on cardiovascular disease, poor health of the respiratory and musculoskeletal systems and its potential damage to the nervous system. We observe an influence of obesity on gastrointestinal disorders, psychiatric phenotypes and the neurodevelopmental disorder *ADHD*. Also, we identified causal associations of obesity on bioelectrical impedances and physical disability. Altogether, our results confirm some previously reported associations and flag out some new testable hypotheses that could contribute to advance our understanding of the effects of obesity on metabolic inflammation in specific tissues and organs, which in turn may provide novel perspectives on the metabolic implications of obesity and the development of anti-inflammatory therapeutics.

## Data Availability

Data can be obtained from the authors upon reasonable request. The results can be perused on CTG-VIEW.

https://view.genoma.io/gwas/5d81f54cbd15223346bab2cd

## DECLARATIONS

### Funding Info

L.M.G.M and A.I.C. are supported by UQ Research Training Scholarships from The University of Queensland (UQ). M.E.R. thanks support of the NHMRC and Australian Research Council (ARC) through a Research Fellowship (GNT1102821). P.F.K. is supported by an Australian Government Research Training Program Scholarship from Queensland University of Technology (QUT).

### Author contribution

M.E.R. and G.C.-P. conceived and directed the study. L.M.G.-M. performed the statistical and bioinformatics analyses, with support and input from A.I.C., P-F.K., N.G.M., G.C.-P. and M.E.R. L.M.G.-M. wrote the first draft of the paper and integrated input and feedback from all co-authors.

### Data Availability

Individual-level data for UK Biobank participants are available to eligible researchers through the UK Biobank (www.biobank.ac.uk).

### Code Availability

Code used as part of the present manuscript’s work is available from the authors upon reasonable request.

### Compliance with ethical standards

#### Ethics Approval

This study was approved by the Human Research Ethics Committee of the QIMR Berghofer Medical Research Institute.

### Animal Research (Ethics)

Not applicable

### Consent to Participate (Ethics)

Informed consent was obtained from all individual participants included in the study.

### Consent to Publish (Ethics)

All participants provided informed consent for the publication of study results.

## ACKNOWLEDGEMENTS

AIC and LMGM are supported by UQ Research Training Scholarships from The University of Queensland (UQ). MER thanks the National Health and Medical Research Council and Australian Research Council’s support through a Research Fellowship (APP1102821).

## SUPPLEMENTARY FILES

**Online Resource 1**. LCV output for obesity (FDR<5%).

**Online Resource 2**. LCV output for obesity (Bonferroni <5%).

## REFERENCES

Anandacoomarasamy A, Caterson I, Sambrook P, et al (2008) The impact of obesity on the musculoskeletal system. Int J Obes 32:211–222. https://doi.org/10.1038/sj.ijo.0803715

Andersen CJ, Murphy KE, Fernandez ML (2016) Impact of Obesity and Metabolic Syndrome on Immunity. Adv Nutr 7:66–75. https://doi.org/10.3945/an.115.010207

Ansa BE, Hoffman Z, Lewis N, et al (2019) Aspirin Use among Adults with Cardiovascular Disease in the United States: Implications for an Intervention Approach. J Clin Med Res 8.: https://doi.org/10.3390/jcm8020264

Apovian CM (2016) Obesity: definition, comorbidities, causes, and burden. Am J Manag Care 22:

Bulik-Sullivan B, Finucane HK, Anttila V, et al (2015a) An atlas of genetic correlations across human diseases and traits. Nat Genet 47:1236–1241. https://doi.org/10.1038/ng.3406

Bulik-Sullivan BK, Loh P-R, Finucane HK, et al (2015b) LD Score regression distinguishes confounding from polygenicity in genome-wide association studies. at Genet 47:291–295. https://doi.org/10.1038/ng.3211

Camilleri M, Malhi H, Acosta A (2017) Gastrointestinal Complications of Obesity. Gastroenterology 152:1656–1670. https://doi.org/10.1053/j.gastro.2016.12.052

Campia U, Tesauro M, Cardillo C (2012) Human obesity and endothelium-dependent responsiveness. Br J Pharmacol 165:561–573. https://doi.org/10.1111/j.1476-5381.2011.01661.x

Chang P, Friedenberg F (2014) Obesity and GERD. Gastroenterol Clin North Am 43:161–173. https://doi.org/10.1016/j.gtc.2013.11.009

Chauvet-Gelinier J-C, Roussot A, Cottenet J, et al (2019) Depression and obesity, data from a national administrative database study: Geographic evidence for an epidemiological overlap. PLoS One 14:e0210507. https://doi.org/10.1371/journal.pone.0210507

Chen S-Y, Feng Z, Yi X (2017) A general introduction to adjustment for multiple comparisons. J Thorac Dis 9:1725–1729. https://doi.org/10.21037/jtd.2017.05.34

Cortese S (2019) The Association between ADHD and Obesity: Intriguing, Progressively More Investigated, but Still Puzzling. Brain Sci 9.: https://doi.org/10.3390/brainsci9100256

Cortese S, Tessari L (2017) Attention-Deficit/Hyperactivity Disorder (ADHD) and Obesity: Update 2016. Curr Psychiatry Rep 19:4. https://doi.org/10.1007/s11920-017-0754-1

Cuéllar-Partida G, Lundberg M, Kho PF, et al (2019) Complex-Traits Genetics Virtual Lab: A community-driven web platform for post-GWAS analyses. bioRxiv 518027

Davis BB, Shen Y-H, Tancredi DJ, et al (2012) Leukocytes are recruited through the bronchial circulation to the lung in a spontaneously hypertensive rat model of COPD. PLoS One 7:e33304. https://doi.org/10.1371/journal.pone.0033304

Day FR, Ong KK, Perry JRB (2018) Elucidating the genetic basis of social interaction and isolation. Nat Commun 9:2457. https://doi.org/10.1038/s41467-018-04930-1

De Lorenzo A, Soldati L, Sarlo F, et al (2016) New obesity classification criteria as a tool for bariatric surgery indication. World J Gastroenterol 22:681–703. https://doi.org/10.3748/wjg.v22.i2.681

Dixon AE, Peters U (2018) The effect of obesity on lung function. Expert Rev Respir Med 12:755– 767. https://doi.org/10.1080/17476348.2018.1506331

García-Marín LM, Campos AI, Martin NG, et al (2021) Inference of causal relationships between sleep-related traits and 1,527 phenotypes using genetic data. Sleep 44.: https://doi.org/10.1093/sleep/zsaa154

Ghesmaty Sangachin M, Cavuoto LA, Wang Y (2018) Use of various obesity measurement and classification methods in occupational safety and health research: a systematic review of the literature. BMC Obes 5:28. https://doi.org/10.1186/s40608-018-0205-5

Goodarzi MO (2018) Genetics of obesity: what genetic association studies have taught us about the biology of obesity and its complications. The Lancet Diabetes & Endocrinology 6:223–236. https://doi.org/10.1016/S2213-8587(17)30200-0

Hawkley LC, Cacioppo JT (2010) Loneliness matters: a theoretical and empirical review of consequences and mechanisms. Ann Behav Med 40:218–227. https://doi.org/10.1007/s12160-010-9210-8

Haworth S, Kho PF, Holgerson PL, et al (2020) Assessment and visualization of phenome-wide causal relationships using genetic data: an application to dental caries and periodontitis. Eur J Hum Genet 1–9. https://doi.org/10.1038/s41431-020-00734-4

Higgins V, Nazroo J, Brown M (2019) Pathways to ethnic differences in obesity: The role of migration, culture and socio-economic position in the UK. SSM Popul Health 7:100394. https://doi.org/10.1016/j.ssmph.2019.100394

Hoffman M, Blum A, Baruch R, et al (2004) Leukocytes and coronary heart disease. Atherosclerosis 172:1–6. https://doi.org/10.1016/S0021-9150(03)00164-3

Hozumi J, Sumitani M, Matsubayashi Y, et al (2016) Relationship between Neuropathic Pain and Obesity. Pain Res Manag 2016:2487924. https://doi.org/10.1155/2016/2487924

Hruby A, Hu FB (2015) The Epidemiology of Obesity: A Big Picture. Pharmacoeconomics 33:673– 689. https://doi.org/10.1007/s40273-014-0243-x

Hurt RT, Kulisek C, Buchanan LA, McClave SA (2010) The obesity epidemic: challenges, health initiatives, and implications for gastroenterologists. Gastroenterol Hepatol 6:780–792

Kaur Y, Wang DX, Liu H-Y, Meyre D (2019) Comprehensive identification of pleiotropic loci for body fat distribution using the NHGRI-EBI Catalog of published genome-wide association studies. Obes Rev 20:385–406. https://doi.org/10.1111/obr.12806

Kim JH, Lim S, Park KS, et al (2017) Total and differential WBC counts are related with coronary artery atherosclerosis and increase the risk for cardiovascular disease in Koreans. PLoS One 12:e0180332. https://doi.org/10.1371/journal.pone.0180332

Koellinger PD, de Vlaming R (2019) Mendelian randomization: the challenge of unobserved environmental confounds. Int J Epidemiol 48:665–671. https://doi.org/10.1093/ije/dyz138

Koo H-K, Kang HK, Song P, et al (2017) Systemic White Blood Cell Count as a Biomarker Associated with Severity of Chronic Obstructive Lung Disease. Tuberc Respir Dis 80:304–310. https://doi.org/10.4046/trd.2017.80.3.304

Langer HF, Chavakis T (2009) Leukocyte-endothelial interactions in inflammation. J Cell Mol Med 13:1211–1220. https://doi.org/10.1111/j.1582-4934.2009.00811.x

Leick M, Azcutia V, Newton G, Luscinskas FW (2014) Leukocyte recruitment in inflammation: basic concepts and new mechanistic insights based on new models and microscopic imaging technologies. Cell Tissue Res 355:647–656. https://doi.org/10.1007/s00441-014-1809-9

Liu C-Y, Schoeler T, Davies NM, et al (2020) Are there causal relationships between attention-deficit/hyperactivity disorder and body mass index? Evidence from multiple genetically informed designs. Int J Epidemiol. https://doi.org/10.1093/ije/dyaa214

Locke AE, Kahali B, Berndt SI, et al (2015) Genetic studies of body mass index yield new insights for obesity biology. Nature 518:197–206. https://doi.org/10.1038/nature14177

Lumeng CN, Saltiel AR (2011) Inflammatory links between obesity and metabolic disease. J Clin Invest 121:2111–2117. https://doi.org/10.1172/JCI57132

Luppino FS, de Wit LM, Bouvy PF, et al (2010) Overweight, obesity, and depression: a systematic review and meta-analysis of longitudinal studies. Arch Gen Psychiatry 67:220–229. https://doi.org/10.1001/archgenpsychiatry.2010.2

Mafort TT, Rufino R, Costa CH, Lopes AJ (2016) Obesity: systemic and pulmonary complications, biochemical abnormalities, and impairment of lung function. Multidiscip Respir Med 11:28. https://doi.org/10.1186/s40248-016-0066-z

Mannan M, Mamun A, Doi S, Clavarino A (2016) Prospective Associations between Depression and Obesity for Adolescent Males and Females-A Systematic Review and Meta-Analysis of Longitudinal Studies. PLoS One 11:e0157240. https://doi.org/10.1371/journal.pone.0157240

Martins-Silva T, dos Santos Vaz J, Hutz MH, et al (2019) Assessing causality in the association between attention-deficit/hyperactivity disorder and obesity: a Mendelian randomization study. Int J Obes 43:2500–2508. https://doi.org/10.1038/s41366-019-0346-8

Mitchell NS, Catenacci VA, Wyatt HR, Hill JO (2011) Obesity: overview of an epidemic. Psychiatr Clin North Am 34:717–732. https://doi.org/10.1016/j.psc.2011.08.005

Neale’s Lab (2018) GWAS Results. In: UK Biobank - Neale Lab. http://www.nealelab.is/uk-biobank. Accessed 29 Jul 2020

Ng M, Fleming T, Robinson M, et al (2014) Global, regional, and national prevalence of overweight and obesity in children and adults during 1980-2013: a systematic analysis for the Global Burden of Disease Study 2013. Lancet 384:766–781. https://doi.org/10.1016/S0140-6736(14)60460-8

Noble WS (2009) How does multiple testing correction work? Nat Biotechnol 27:1135–1137. https://doi.org/10.1038/nbt1209-1135

O’Connor LJ, Price AL (2018) Distinguishing genetic correlation from causation across 52 diseases and complex traits. Nat Genet 50:1728–1734. https://doi.org/10.1038/s41588-018-0255-0

Panuganti KK, Nguyen M, Kshirsagar RK (2020) Obesity. In: StatPearls [Internet]. StatPearls Publishing

Purnell JQ (2018) Definitions, Classification, and Epidemiology of Obesity. In: Endotext [Internet]. MDText.com, Inc.

Queirós FC, Wehby GL, Halpern CT (2015) Developmental disabilities and socioeconomic outcomes in young adulthood. Public Health Rep 130:213–221. https://doi.org/10.1177/003335491513000308

Skyler JS, Bakris GL, Bonifacio E, et al (2017) Differentiation of Diabetes by Pathophysiology, Natural History, and Prognosis. Diabetes 66:241–255. https://doi.org/10.2337/db16-0806

Speed MS, Jefsen OH, Børglum AD, et al (2019) Investigating the association between body fat and depression via Mendelian randomization. Transl Psychiatry 9:184. https://doi.org/10.1038/s41398-019-0516-4

Stevens JR, Al Masud A, Suyundikov A (2017) A comparison of multiple testing adjustment methods with block-correlation positively-dependent tests. PLoS One 12:e0176124. https://doi.org/10.1371/journal.pone.0176124

Swarup S, Goyal A, Grigorova Y, Zeltser R (2020) Metabolic Syndrome. In: StatPearls [Internet]. StatPearls Publishing

Tam V, Turcotte M, Meyre D (2019) Established and emerging strategies to crack the genetic code of obesity. Obes Rev 20:212–240. https://doi.org/10.1111/obr.12770

Yengo L, Sidorenko J, Kemper KE, et al (2018) Meta-analysis of genome-wide association studies for height and body mass index in □700000 individuals of European ancestry. Hum Mol Genet 27:3641–3649. https://doi.org/10.1093/hmg/ddy271

Zammit C, Liddicoat H, Moonsie I, Makker H (2010) Obesity and respiratory diseases. Int J Gen Med 3:335–343. https://doi.org/10.2147/IJGM.S11926

